# The β-NGF/TrkA signalling pathway is associated with the production of anti- nucleoprotein IgG in convalescent COVID-19

**DOI:** 10.1101/2021.11.11.21266223

**Authors:** Carla Usai, Joseph M. Gibbons, Corinna Pade, Wenhao Li, Sabina R.M. Jacobs, Áine McKnight, Patrick T. F. Kennedy, Upkar S. Gill

**Author notes:** Joint senior & corresponding authors **Correspondence:** Dr Upkar S. Gill & Professor Patrick T.F. Kennedy. These authors have contributed equally to this work (Joseph M. Gibbons, Corinna Pade).

## Abstract

**Background:** The presentation of SARS-CoV-2 infection varies from asymptomatic to severe COVID. Similarly, high variability in the presence, titre and duration of specific antibodies has been reported. While some host factors determining these differences, such as age and ethnicity have been identified, the underlying molecular mechanisms underpinning these differences remain poorly defined.

**Methods:** We analysed serum and PBMC from 17 subjects with a previous PCR confirmed SARS-CoV-2 infection and 10 unexposed volunteers following the first wave of the pandemic, in the UK. Anti-NP IgG and neutralising antibodies were measured, as well as a panel of infection and inflammation related cytokines. The virus-specific T cell response was determined by IFN-γ ELISPOT and flow cytometry after over-night incubation of PBMCs with pools of selected SARS-CoV-2 specific peptides.

**Results:** Seven of 17 convalescent subjects had undetectable levels of anti-NP IgG, and a positive correlation was shown between anti-NP IgG levels and the titre of neutralising antibodies (IC50). In contrast, a discrepancy was noted between antibody levels and T cell IFN-γ production by ELISpot following stimulation with specific peptides. Among the analysed cytokines, β-NGF and IL-1α levels were significantly different between anti-NP positive and negative subjects, and only β-NGF significantly correlated with anti-NP positivity. Interestingly, CD4+ T cells of anti-NP negative subjects expressed lower amounts of the β-NGF-specific receptor TrkA.

**Conclusions:** Our results suggest that the β-NGF/TrkA signalling pathway is associated with the production of anti-NP specific antibody in mild SARS-CoV-2 infection and the mechanistic regulation of this pathway in COVID-19 requires further investigation.

## Introduction

SARS-CoV-2 infected subjects can display symptoms within a wide range of severity, from asymptomatic or pauci-symptomatic forms (characterised by fever, cough, fatigue, sore throat, loss of smell) to respiratory failure and systemic manifestations (sepsis, septic shock, and multiple organ dysfunction syndromes (1). Similarly, high variability in the presence, titre and duration of specific antibodies has been reported (2–4), often positively correlating with disease severity (3–5).

Some factors determining differences in clinical manifestations and humoral response, such as age, ethnicity and co- or pre-existing medical conditions have already been described (2,6). Underlying genetic and molecular determinants of humoral responses are currently being investigated with some promising results, although these studies mainly focussed on subjects experiencing severe COVID-19 (7–9).

It is estimated that 90-99% of PCR confirmed SARS-CoV-2 infected individuals mount a specific humoral response, while 1-10% have very low to undetectable anti-spike (S) or anti-nucleoprotein (NP) IgG by commercial serological assays (4,10–12). Likewise, specific SARS-CoV-2 T cells have been detected in the majority of COVID-19 convalescent patients, even in cases where humoral responses are undetectable (13,14).

While the presence of IgG against S and NP of SARS-CoV-2 are known to be detected with varying kinetics (3,15), T cell responses appear to be simultaneously directed to several antigens from early phases of SARS-CoV-2 exposure (16,17). The possibility of an existing hierarchy with some of the viral antigens being more efficient in eliciting a T cell response, or because of cross-reactive T cells due to previous infections in some individuals has been considered (5,18,19).

It has been described that an early T cell response during active SARS-CoV-2 infection is associated with milder symptoms and rapid viral clearance (5). Regarding the humoral response, associations between anti-NP and anti-S dominated early responses with different outcomes have been found in independent studies, with severe COVID-19 patients showing an early response dominated by anti-NP antibodies, and mild to moderate cases exhibiting a dominant anti-S response (5,20,21). A better characterisation of such inter-individual variability identifying prognostic factors will allow better stratification according to the relative risk of developing severe disease, which may be key to prioritise future treatment and vaccination strategies.

To address this question, we utilised a cohort of subjects sampled immediately following the first wave of the COVID-19 pandemic in the UK; we analysed serum samples and peripheral mononuclear cells (PBMC) from 7 anti-NP negative, 10 anti-NP positive COVID-19 convalescent subjects, and 10 unexposed volunteers. We determined the titres of neutralising antibodies, the presence of antigen-specific T cells, and serum levels of cytokines related to infection and inflammation, to identify host factors associated with anti-NP IgG positivity. We identify an association between the presence of circulating anti-NP antibodies and the nerve growth factor (β-NGF)/TrkA pathway, known to be active in lymphocytes and to be involved in inflammatory conditions of the airways (22–25).

## Material and Methods

### Convalescent COVID-19 and Healthy Donors

Forty donors were randomly selected from a previously published cohort (2) to create four sex- and age-matched groups according to PCR and antibody status. Group 1: negative PCR and negative anti-NP IgG n=10 (“unexposed”); Group 2: positive PCR and positive anti-NP IgG n=10; Group 3: positive PCR and negative anti-NP IgG n=7; negative or n/a PCR and positive anti-NP IgG n=13. All participants provided informed consent according to the local ethics committee approval (Approved 22/04/2020, South Central - Berkshire Research Ethics Committee ref: 20/SC/0191, ISRCTN60400862).

### Sample Collection

Venepuncture was performed on each participant utilising the sites standard blood collection method. 40 ml of whole blood were collected in EDTA plasma vacutainers for serum collection and lithium heparin tubes for peripheral blood cell isolation. Serum samples were obtained by centrifugation of 5 ml venous blood at room temperature at 3,000 g for 15min, aliquoted and frozen on the day of collection.

### PBMC isolation

PBMCs were isolated from heparinised blood by density gradient centrifugation on Ficoll-Paque (2000 rpm for 23 minutes at room temperature with minimum deceleration speed) and cells immediately frozen in fetal bovine serum 10% DMSO. Cells were thawed on the day of experimentation and used directly for the *ex vivo* experiments.

### Antibody tests

The presence of anti-Nucleocapsid protein (NP) IgG and IgM in serum samples was determined using the Panbio™ COVID-19 IgG/IgM Rapid Test Device (Fingerstick Whole Blood/Venous Whole Blood/Serum/Plasma) (Panbio™; Abbott Rapid Diagnostics Jena GmbH, Jena, Germany) according to the manufacturer’s instructions and as previously described (2). Anti-NPIgG leves in serum samples were quantified using the Abbott Architect i2000 chemiluminescent microparticle immunoassay (Architect) was for SARS-CoV-2 IgG (Abbott Diagnostics, IL, USA; Architect) according to the manufacturer’s instructions and as previously described (2).

### Authentic Virus Neutralisation Assay

SARS-CoV-2 microneutralisation assay was performed as described previously (14,26). VeroE6 cells were seeded in 96-well plates 24h prior to infection. Duplicate titrations of heat-inactivated patient sera were incubated with 3×104 FFU SARS-CoV-2 virus (TCID100) at 37°C for 1h. Serum/virus preparations were added to cells and incubated for 72h. Surviving cells were fixed in 3.7% (vol/vol) formaldehyde and stained with 0.1% (wt/vol) crystal violet solution. Crystal violet stain was resolubilised in 1% (wt/vol) sodium dodecyl sulphate solution. Absorbance readings were taken at 570nm using a CLARIOStar Plate Reader (BMG Labtech). Negative controls of pooled pre-pandemic sera (collected prior to 2019), and pooled serum from neutralisation positive SARS-CoV-2 convalescent individuals were spaced across the plates. Absorbance for each well was standardised against technical positive (virus control) and negative (cells only) controls on each plate to determine percentage neutralisation values. IC50s were determined from neutralisation curves. All authentic SARS-CoV-2 propagation and microneutralisation assays were performed in a containment level 3 facility.

### Cytokine analysis

Serum levels of a customised panel of cytokines and chemokines were determined using cytokine bead assay (CBA) kits (Bio-Techne Ltd) on a Magpix (Luminex Corporation) equipped with xPonent® software for data acquisition and analysis. According to the manufacturers instruction, serum samples were diluted at a ratio of 1:2 for the determination of Leptin, CCL2, GM-CSF, HGF, IFN-γ, IL-1α, IL-1β, IL-6, IL-8, IL-10, IL-12 p70, β-NGF, PBEF/Visfatin, Resistin, PAI 1, TNF-α and VEGF concentration; 1:200 for Adiponectin, Serpin A12 and C reactive protein, and 1:400 for RBP4 analysis.

### -CoV-2 peptide pools

Twenty peptides from the Spike, Membrane, Nucleoprotein and ORF-7a-2 proteins of SARS-CoV-2 containing T cell epitopes with known immunogenic properties (17,27) were purchased from ProImmune Limited. The purity of the peptides was above 80%, and their composition was confirmed by mass spectrometry analysis. Single peptides were reconstituted in DMSO and pooled as outlined in **Table 1** and used at a final concentration of 2 µM, reconstituted in RPMI.

**Table 1.**
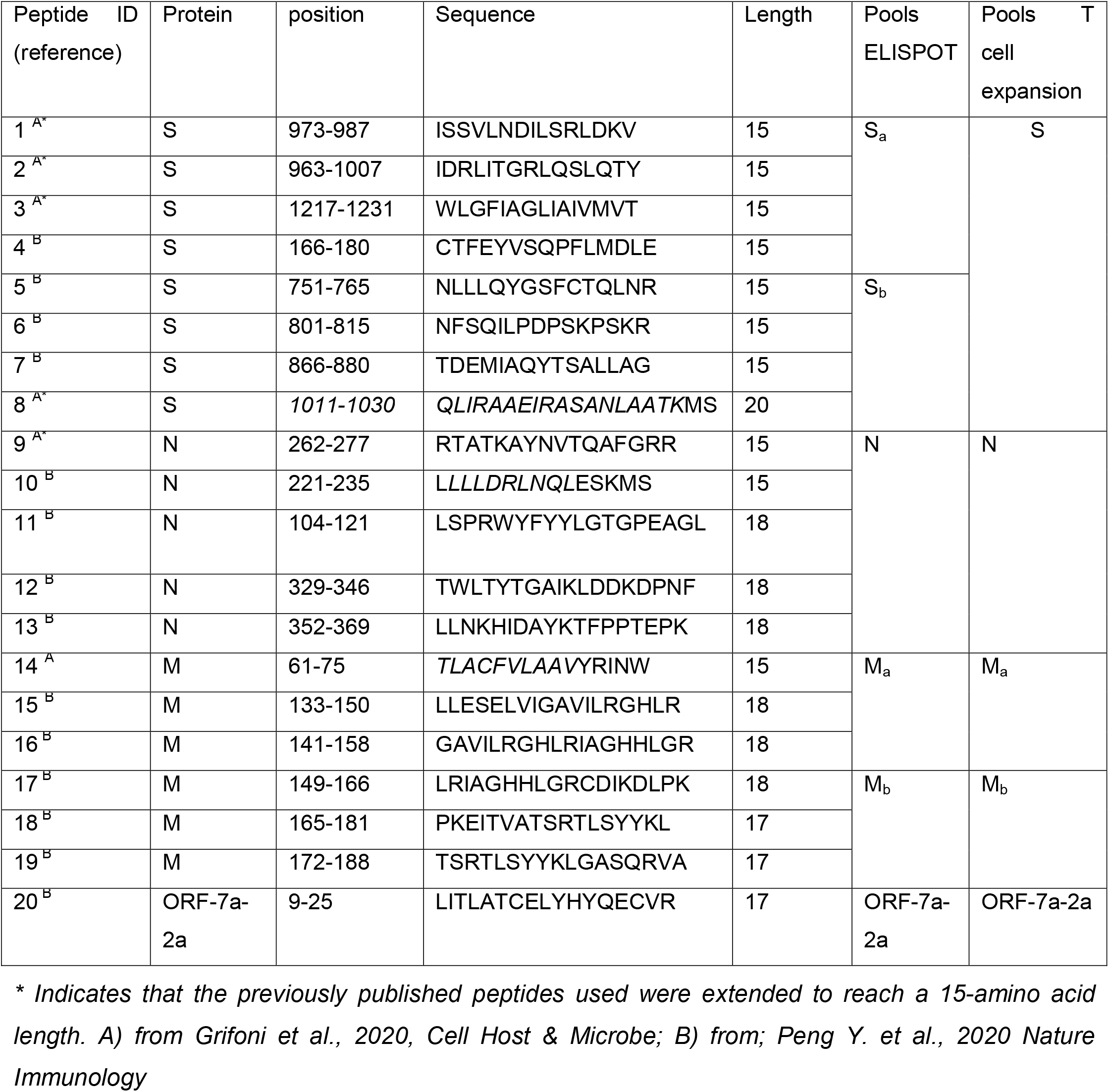
SARS-CoV-2 peptide pools used for IFN-γ ELISpot assay and T cell expansion experiments.

### IFN-γ ELISPOT assay

IFN-γ ELISpot assays were performed with cryopreserved PBMCs, using human IFN-γ ELISPOT Set (BD, 551849). PBMC were thawed, washed twice in RPMI media, and seeded at a final concentration of 2 × 10^5^ cells/100 µl and stimulated for 18 hours with 100 µl/well of each peptide pool at a final concentration of 1µM/peptide/well, at 37°C, 5% CO_2_. Treatment with PMA (Abcam, ab120297) and Ionomycin (Abcam ab120370) (final concentration 250 ng/ 5 µg/ ml respectively) was used as a positive control, while RPMI was added to unstimulated cells. Spot forming units (SFU) were quantified with a BIOREADER® 7000–F (BioSys GmbH). To quantify positive peptide-specific responses, 2× mean spots of the unstimulated wells were subtracted from the peptide-stimulated wells, and the results expressed as SFU/10^6^ cells.

### T cell expansion

Frozen PBMCs were thawed and washed twice in 5 ml RPMI medium, centrifuged at 1500 rpm for 5 min and resuspended at a final concentration of 2 × 10^6^/ml in RPMI 10%FBS 2 µM Monensin (eBioscience, 00-4505-51). 100 µl cell suspensions were stimulated in a 96-well plate for 18 hours with 100 µl/well of each peptide pool, at a final concentration of 1µM/peptide/well, at 37°C, 5% CO_2_. Treatment with PMA (Abcam, ab120297) and Ionomycin (Abcam ab120370) at the final concentration of 50 ng/ml and 1 µg/ml respectively, was used as a positive control, while RPMI was added to unstimulated cells. Cells were washed in 100 µl PBS 1x. Each well was incubated for 20 min at 4°C in the dark, with saturating concentrations (100 µl) of a mix of the following antibodies: anti-PD1 PE-Cy7 (BioLegend, clone EH12.2H7), anti-CD8 APC-Cy7 (BioLegend, clone SK1), anti-TrkA PE (R&D Systems), anti-CD3 V500 (BD Biosciences, clone UCHT1), anti-CD4 BV605 (BioLegend, clone OKT4), anti-CD69 AF700 (BioLegend, clone FN50), anti-KLRG1 PerCP (BioLegend). The Blue Fluorescent Reactive Dye (Invitrogen, L23105) was added to the mix to assess cell viability. Cells were subsequently fixed and permeabilized using the Cytofix/Cytoperm kit (BD Biosciences — Pharmingen) (100 µl/well for 30 min at 4°C in the dark) and stained with anti– IFN-γ BV450 (BD Biosciences, clone B27), anti–TNF-α APC (BioLegend, clone Mab11), anti–IL-2 PE-CF594 (clone 5344.111, BD Horizon), anti-MIP-1β FITC (clone D21-12351 BD Pharmingen). Cells were acquired on a BD-LSR II FACS Scan, and data were analysed using FlowJo (Tree Star Inc.).

### Statistical analysis

Statistical analysis was performed using GraphPad Prism 9.1.2, GraphPad Software, San Diego, California USA, (www.graphpad.com). Specific statistical tests for each experiment are included in the representative figure legends; p values <0.05 were considered significant. Binary logistic regression was performed using IBM SPSS Statistics for Windows, Version 27. Armonk, NY: IBM Corp.

## Results

### Discordant antibody levels relative to T cell responses

We have previously determined the anti-NP IgM and IgG levels of 228 volunteers, after the first wave of the pandemic in the United Kingdom (2). Seven out of 87 participants who had had a positive PCR test, had undetectable levels of anti-NP IgG, irrespective of symptomatology (**Table 2**). Forty donors were randomly selected into four sex- and age-matched groups (**Supplementary Table 1**), and their T cell response to selected peptide pools was analysed with IFN-γ ELISPOT (of the 7 PCR positive, anti-NP IgG negative subject, only 4 had given their consent for additional PBMC isolation).

**Table 2.** Number of subjects shown from entire (2)

|  | N | Anti-NP IgG positive (N) | Anti-NP IgG Negative (N) |
| --- | --- | --- | --- |
| **PCR positive** | **87** | **80** | **7** |
| Symptomatic | 71 | 67 | 4 |
| Asymptomatic | 16 | 13 | 3 |
| **PCR negative** | **23** | **3** | **20** |
| Symptomatic | 4 | 0 | 4 |
| Asymptomatic | 19 | 3 | 16 |
| **PCR N/A** | **118** | **20** | **98** |
| Symptomatic | 12 | 6 | 6 |
| Asymptomatic | 106 | 14 | 92 |
| **Total** | **228** | **103** | **125** |

A large proportion (80%) of subjects who were PCR negative, anti-NP IgG negative did not produce IFN-γ following SARS-CoV-2 peptide stimulation, whereas 83% of those demonstrating PCR positivity, regardless of anti-NP IgG status, demonstrated positive ELISPOT responses as marked by IFN-γ spot-forming units (SFUs). In subjects where a PCR result was not available, 7 of 9 (78%) demonstrated evidence of IFN-γ production after overnight stimulation with the selected peptide pools, with a proportion of responders close to that of the PCR positive anti-NP positive group (7 out of 8, 87.5%) (**Figure 1A**). Deconvoluting the total T cell responses into individual peptide pools, derived from four SARS-CoV-2 proteins (Membrane – M, Nucleoprotein –NP, ORF-7a-2, and Spike –S), we observed a similar number of responders with a similar distribution among the groups. Surprisingly, however those PCR positive anti-NP IgG negative subjects did not produce recordable IFN-γ responses following stimulation with the pool of peptides derived from S (**Figure 1B**). Since we considered the PCR status as one of the defining characteristics of our subject cohorts, we elected to exclude the subjects where PCR results were not available from subsequent analyses.

**Figure 1.**
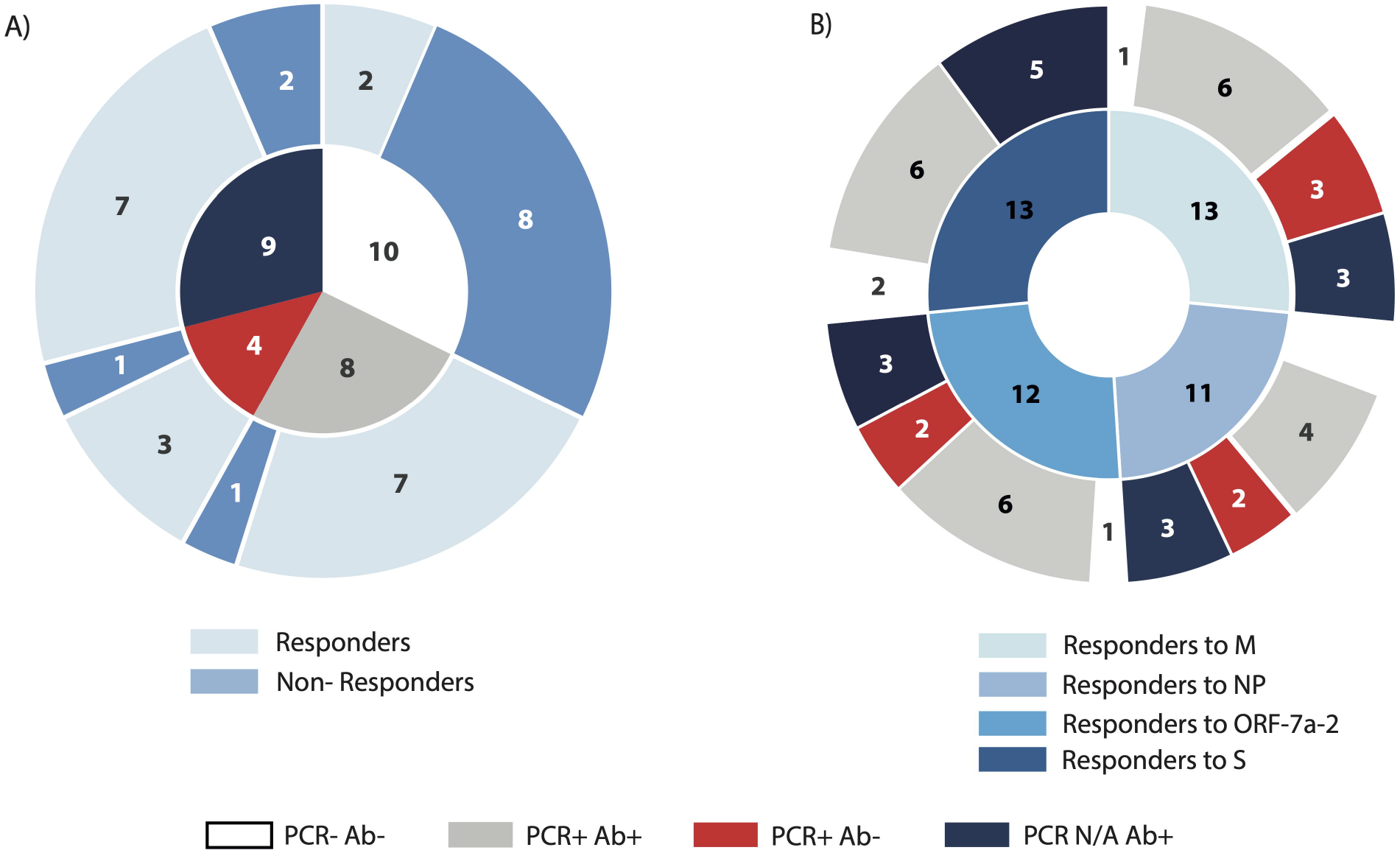
Demonstration of SARS-CoV-2 specific IFN-γ ELISpot responses in relation to PCR and anti-NP status. PBMCs from convalescent COVID-19 subjects and unexposed volunteers were incubated for 16 hours with peptide pools derived from four different SARS-CoV-2 proteins (M: membrane; NP: nucleoprotein; ORF-7a-2: open reading frame 7a-2; S: spike). **(A)** The inner circle represents the composition of the cohort according to PCR and anti-NP (antibody) status; the outer circle represents the proportion of subjects producing IFN-γ (Responders) or absent of IFN-γ production (Non-Responders), following incubation with the total SARS-CoV-2 peptide pool. **(B)** The inner circle represents the number of samples in the cohort producing IFN-γ following incubation with each of the peptide pools indicated; the outer circle represents the distribution of the IFN-γ-producing samples across the four characterised groups according to PCR and antibody status. N/A: not available.

In addition to the Abbott Architect and Panbio assay for anti-NP IgG measurement, we determined the presence and titre of neutralising antibodies (nAbs) which has the receptor binding domain (RBD) of the S protein as their major target. In all subjects where anti-NP IgG positivity was demonstrated, high nAb titres (IC50>200) were noted. Neutralising antibody titres were absent (IC50<50), as expected, in all PCR negative subjects but also in two of seven PCR positive subjects with negative anti-NP IgG status. In the five subjects where, anti-NP was negative but nAbs were detectable, two displayed low titres (IC50=50-199) and three high titres (**Figures 2A and 2B, Supplementary Figure 1A and 1B**). While a positive correlation was found between the level of anti-NP IgG and the titre of nAbs (**Supplementary Figure 1C**), there was discordance between antibody levels (either anti-NP or nAbs) and the cumulative T cell response expressed in SFU per million cells (**Figure 2C**), similar to that previously reported (14).

**Figure 2.**
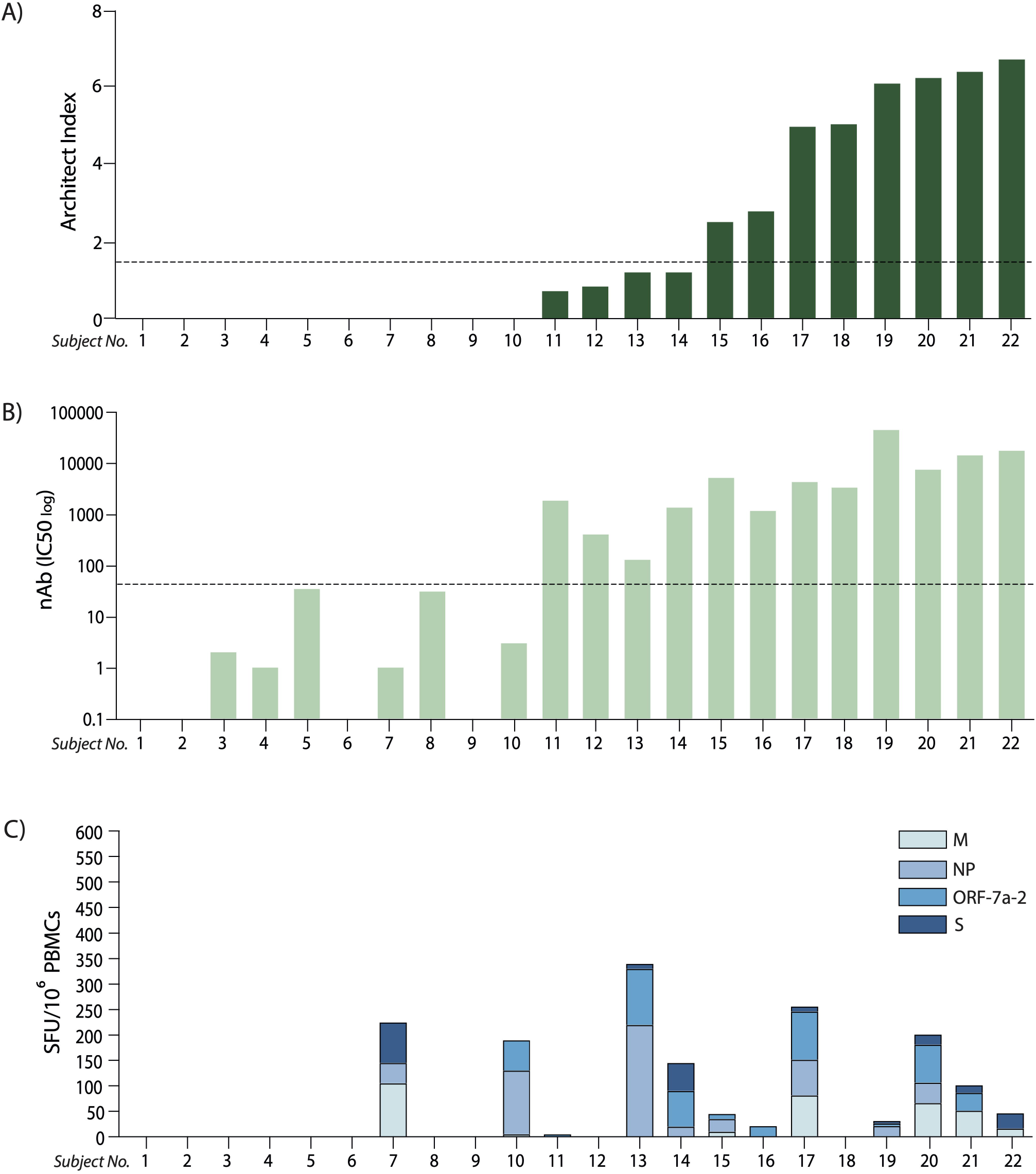
Discordant antibody (anti-NP and nAb) levels with IFN-γ ELISpot responses relative to T cell antigens. **(A)** Anti-nucleoprotein IgG levels expressed as Architect Index (manufacturer arbitrary units) for each subject analysed (ordered lowest to highest level); the dotted line represents the 1.4 cut-off, below which samples are considered as negatives. **(B)** Neutralising antibody (nAb) titres (IC50) corresponding to the same subjects in (A); the dotted line represents the cut-off below which samples are considered as negatives (IC50<50). **(C)** Cumulative T cell response to the four peptide pools derived from SARS-CoV-2 expressed as spot-forming units (SFU) of IFN-γ-secreting cells after 16-hour stimulation, ordered corresponding to the subjects in (A) and (B). (M: membrane; NP: nucleoprotein; ORF-7a-2: open reading frame 7a-2; S: spike).

### Multi-specific and differential CD4 and CD8 T cell cytokine responses according to PCR and antibody status

We then wanted to determine the presence of antigen-specific T cell populations in the peripheral blood of the donors. 2 × 105 PBMCs were stimulated overnight with four peptide pools derived from SARS-CoV-2 and analysed by flow cytometry for the production of IL-2, TNF-α, IFN-γ, and MIP-1β (**Supplementary Figure 2**).

The peptide stimulation following incubation induced the expansion of a small percentage of antigen-specific CD4+ T cells (<1% on average for IL-2, TNF-α and MIP-1β, and <5% for IFN-γ); the frequency of IFN-y producing S-specific cells was higher in PCR positive anti-NP negative subjects compared to unexposed volunteers, and no other significant difference was detected between groups (**Figure 3A**). Among the studied cytokines from CD4+ T cells, TNF-α was produced by the highest proportion of individuals across all groups, ranging from 25% to 100% of subjects depending on the group and the peptide pool used for stimulation, while IL-2 was produced by the lowest proportion of subjects (11%-50%) (**Figure 3C**).

**Figure 3.**
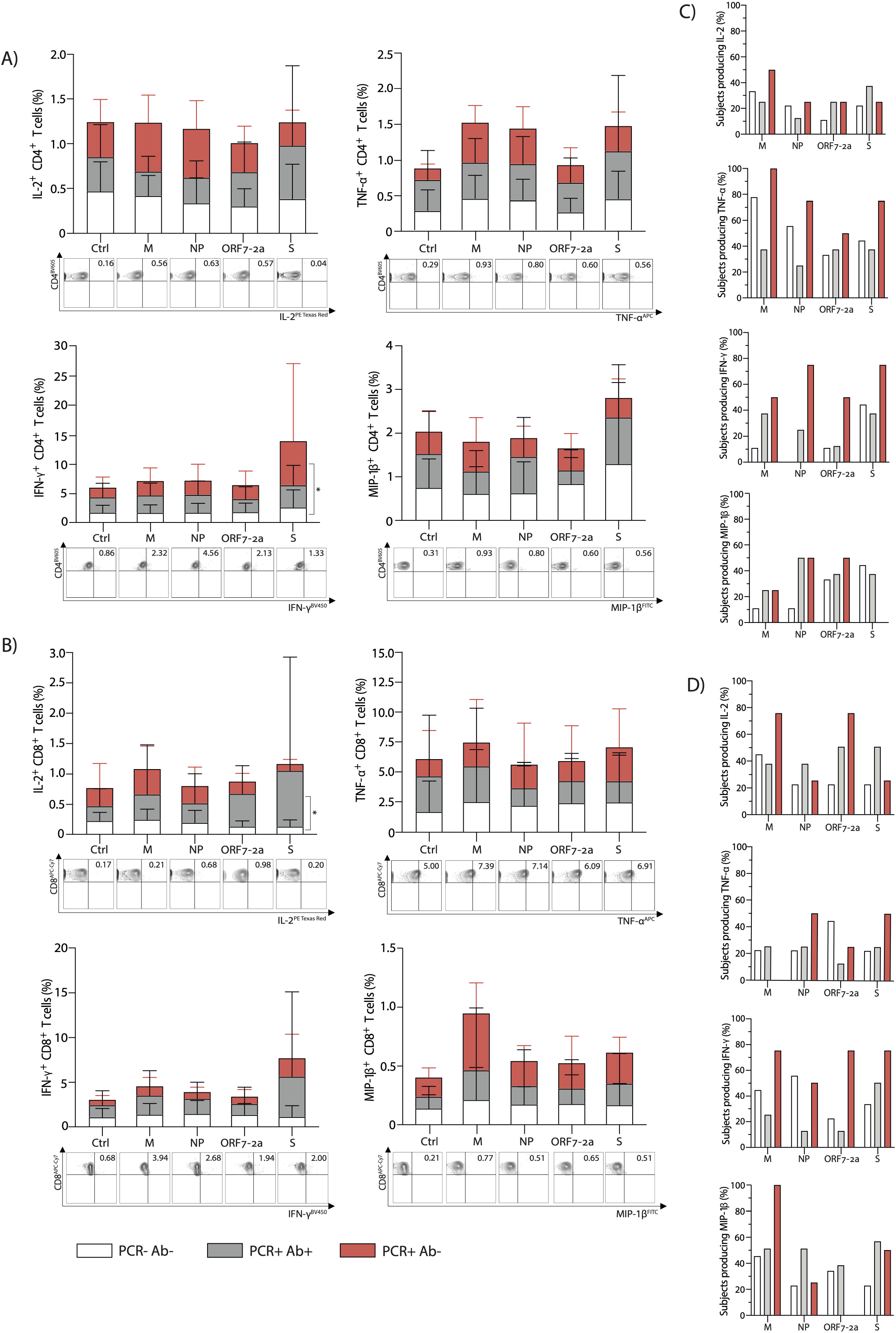
Multi-specific CD4 and CD8 T cell intracellular cytokine responses subsequent to peptide stimulation characterised according to PCR and antibody status. Percentage of **(A)** CD4^+^ and **(B)** CD8^+^ T cells respectively, producing IL-2, TNF-α, IFN-γ, and MIP-1β after 16-hour stimulation with selected peptide pools derived from SARS-CoV-2 (top); representative FACS plots stimulated with the corresponding peptide pools (bottom). Ctrl: cells were incubated with RPMI as a negative control. Percentage of subjects where **(C)** CD4^+^ and **(D)** CD8^+^ T cells produce the cytokines IL-2, TNF-α, IFN-γ, and MIP-1β after 16-hour stimulation with selected peptide pools within each group. (M: membrane; NP: nucleoprotein; ORF-7a-2: open reading frame 7a-2; S: spike). p-values determined by a two–way ANOVA with a Tukey’s post-hoc test for multiple comparisons. *p<0.05; **p<0.01; ***p<0.001, ****p<0.0001. (PCR-Ab- n=10; PCR+Ab+ N=10; PCR+Ab- n=7).

Cytokine production from antigen-specific CD8+ T cells was similarly low when examining IL-2, MIP-1β and IFN-γ (<1% on average for IL-2 and MIP-1β, <5% for IFN-γ), but higher for TNF-α producing cells (up to 4% on average). Moreover, only IL-2-producing S-specific CD8+ T cells were present at a higher frequency in PCR positive anti-NP positive subjects than unexposed controls and no other significant differences were detected (**Figure 3B**). MIP-1β was the cytokine produced by the highest proportion of individuals across groups, ranging from 0% to 100%, depending on the group and the peptide pool used for stimulation, followed by IL-2 and IFN-y (20% and 11% respectively, up to 75%), while TNF-α from CD8+ T cells was produced by lower proportions of subjects (0% to 50%) (**Figure 3D**).

However, when considering the cumulative response of cytokine production to the peptide pools, differences between groups were identified. Within the PCR positive anti-NP positive group the strongest cumulative cytokine production from CD4+ T cells was noted in response to the pool derived from the S protein, with 12.5% of the subjects producing all analysed cytokines; all cytokines were produced with the same frequency in response to this pool. This was followed, in decreasing order, by the pools derived from M (TNF-α and IFN-γ being the predominant cytokines), NP (which induced MIP-1β in 50% of cases), and ORF-7a-2 (mainly inducing TNF-α and MIP-1β). On the contrary, within the PCR positive anti-NP negative group, the greatest cytokine production was achieved in response to the pool derived from the M protein (25% of subjects producing all cytokines, and all producing TNF-α). This was followed by the pools derived from NP (mainly inducing TNF-α) and ORF-7a-2 (inducing the production of TNF-α, IFN-γ and MIP-1β with the same frequencies). The peptide pool derived from S elicited the lowest cumulative cytokine production, dominated by TNF-α and IFN-γ (**Figure 4A**).

**Figure 4.**
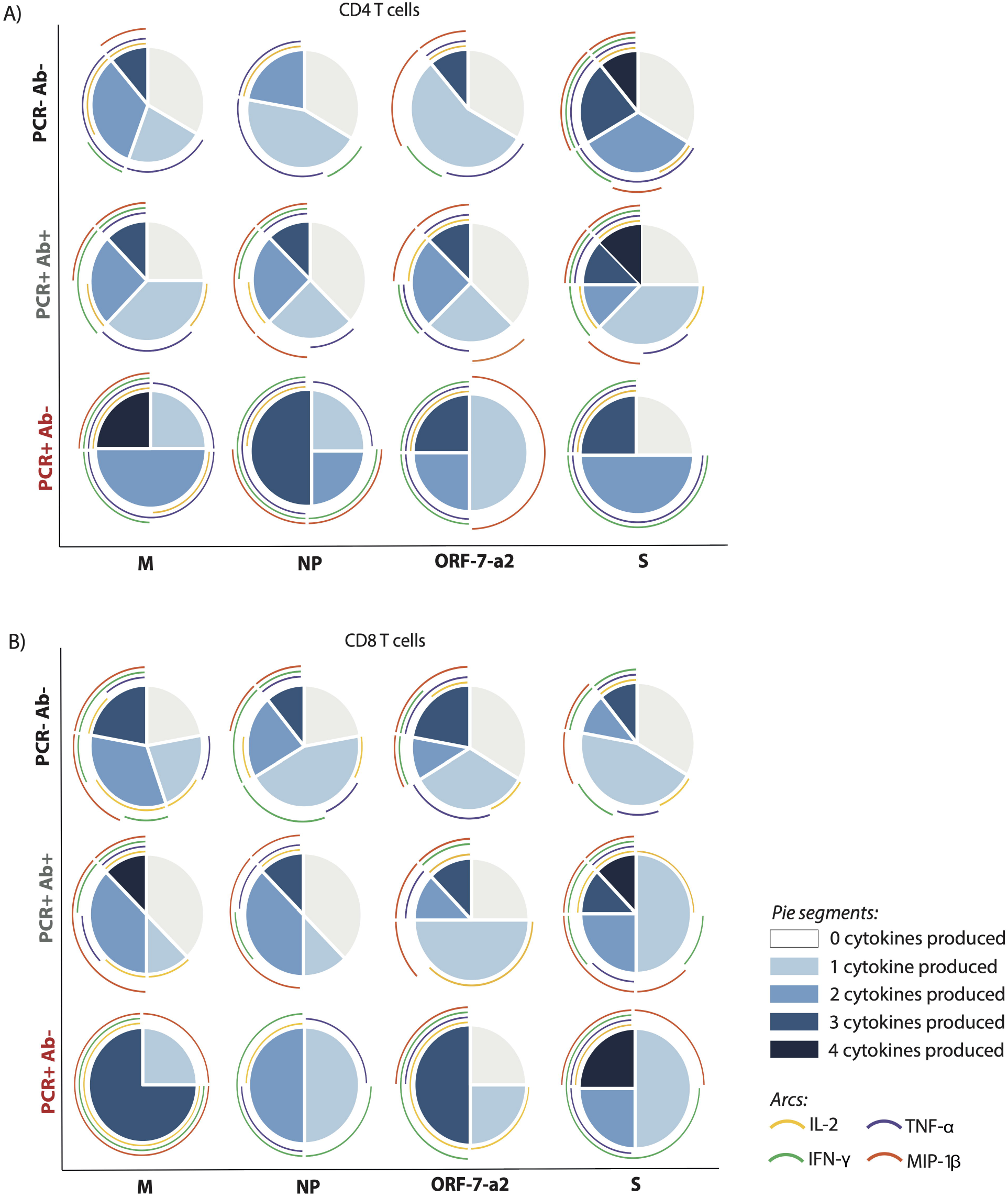
Differential T cells responses, pursuant to the number and type of cytokine produced within the different cohorts. **(A)** CD4^+^ and **(B)** CD8^+^ T cells producing cytokines after stimulation with the four peptide pools derived from SARs-CoV-2. The pie charts represent the proportion of subjects producing a different number of cytokines per group in response to each of the four peptide pools; the arcs show the production of each analysed cytokine, each corresponding to a different colour as indicated. (M: membrane; NP: nucleoprotein; ORF-7a-2: open reading frame 7a-2; S: spike). (PCR-Ab- n=9; PCR+Ab+ n=8; PCR+Ab- n=4).

A similar pattern was observed for CD8+ T cells from the PCR positive, anti-NP positive subjects, with S inducing the strongest cumulative cytokine production (12.5% of subjects producing four cytokines, all of them producing at least one cytokine, and MIP-1β being expressed with the highest frequency). For the remaining pools, 25-37.5% of subjects failed to produce any cytokine; the response was dominated by MIP-1β in the case of the pools derived from M and NP, and by MIP-1β and IL-2 in the case of the peptide derived from ORF-7a-2. However, CD8+ T cells from 25% of the PCR positive anti-NP negative subjects were also able to produce four cytokines in response to the S pool. The lowest cytokine response was elicited by the peptide pool derived from NP, which induced the production of only one or two cytokines per subject sample (TNF-α was induced in three out of four subjects). The pools derived from M and ORF-7a-2 were able to elicit the production of up to three cytokines in two and three subjects out of four respectively, but while the response to M was dominated by MIP-1β, the cytokine induced at the highest frequency in response to ORF-7a-2 was IL-2 (**Figure 4B**). These results suggest that a differential hierarchy of response exists among PCR positive subjects, according to their ability to produce anti-NP antibodies.

### Distinct serum cytokine profiles in relation to anti-NP status

In order to understand whether the differences in the antibody production and T cell response reflected different cytokine profiles, we analysed the levels of 20 infection and inflammation related cytokines in the serum of the selected subjects (**Figure 5A**). GM-CSF, β-NGF, IL-1α, PBEF/Visfatin and IL-12 p70 were found to positively correlate with anti-NP IgG levels (**Figure 5A and 5D, Table 3**), and among them, β-NGF and IL-1α levels were significantly different between PCR positive, anti-NP positive and PCR positive, anti-NP negative subjects (**Figure 5B and 5C**). No correlation was found between serum β-NGF levels and IC50 (**Figure 5E**). A binary logistic regression analysis performed considering anti-NP positivity as a binary variable, confirmed that only β-NGF levels directly correlate with the presence of anti-NP IgG (OR: 11.038, p value=0.010) (**Table 4**).

**Table 3.** Pearson correlation test between the Architect Index and serum cytokines levels.

| Cytokine (pg/ml) | r | P value |
| --- | --- | --- |
| GM-CSF | 0.612 | 0.0007 |
| β-NGF | 0.5954 | 0.0011 |
| IL-1α | 0.5164 | 0.0058 |
| PBEF/Visfatin | 0.4677 | 0.0139 |
| IL-12 p70 | 0.3992 | 0.0391 |

**Table 4.** Binary logistic regression between anti-NP IgG positivity and cytokines levels.

|  |  | Intercept | Standard Error | Wald chi-square test | Degrees of Freedom | P value | Odd ratio |
| --- | --- | --- | --- | --- | --- | --- | --- |
| Step 1<sup>a</sup> | β-NGF (pg/ml) | 2.401 | 0.929 | 6.679 | 1 | 0.010 | 11.038 |
|  | Constant | -6.594 | 2.430 | 7.364 | 1 | 0.007 | 0.001 |
| a. Variable(s) entered on step 1: beta-NGF. |  |  |  |  |  |  |  |

**Figure 5.**
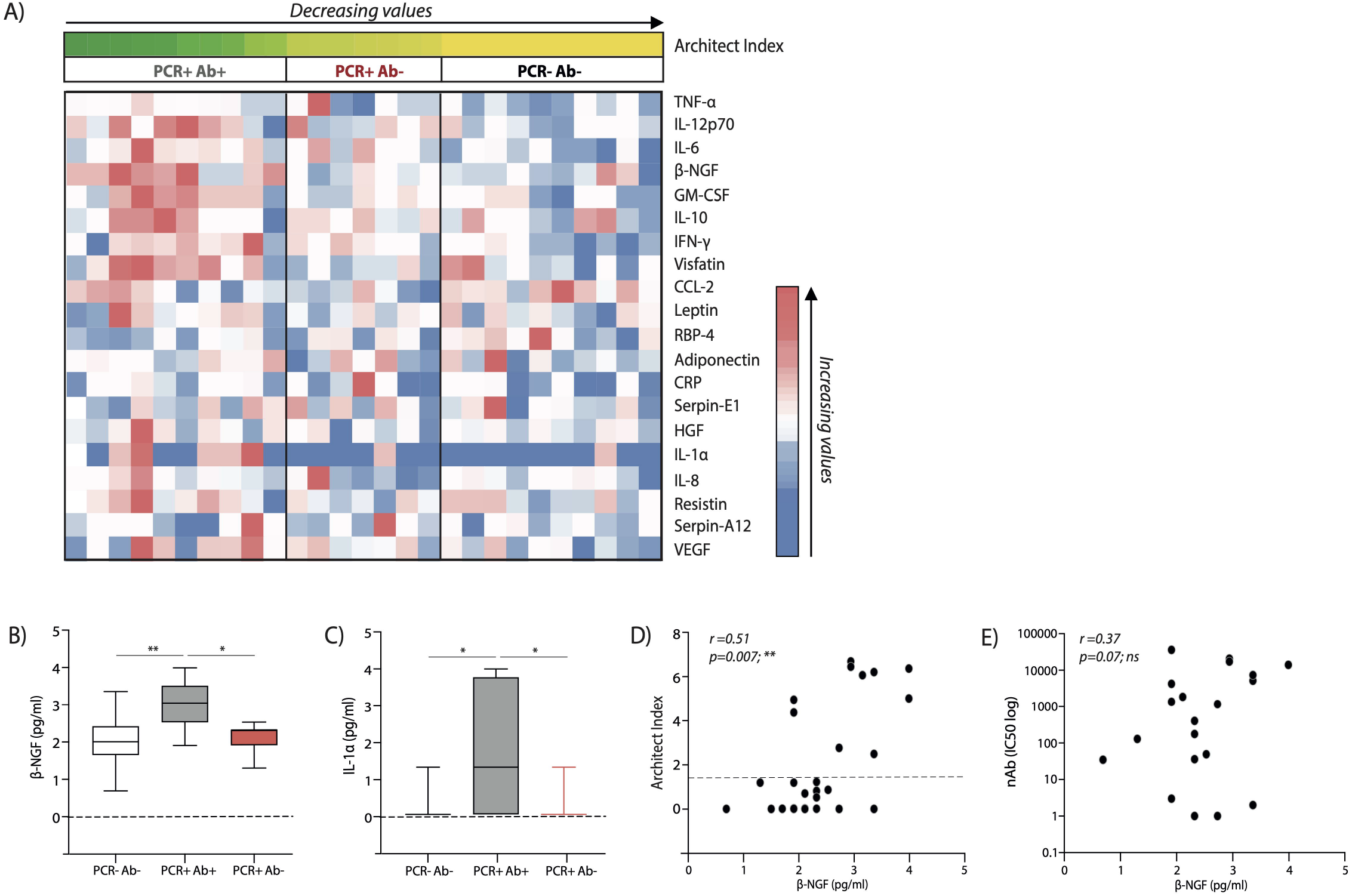
Distinct serum cytokine profiles in the subject cohorts and relative to antibody status. **(A)** Heatmap showing differential serum cytokine expression in the study subjects listed accordingly to respective anti-nucleoprotein IgG levels (Architect Index). **(B)** β-NGF and **(C)** IL-1α serum levels within the 3 different subject cohorts; boxes extend from the 25th to 75th percentiles, the line indicates the median, and the whiskers extend from the smaller to the largest values. Correlative expression of β-NGF with respect to **(D)** Architect index and **(E)** nAb levels, within the whole cohort. p-values in (B) and (C) were determined by a one-way ANOVA with a Tukey’s post-hoc test for multiple comparisons. A Spearman non-parametric correlation test was undertaken to test significance in (D) and (E). *p<0.05; **p<0.01; ***p<0.001; ****p<0.0001; ns - not significant. (PCR-Ab- n=10; PCR+Ab+ N=10; PCR+Ab- n=7).

### T cells from anti-NP IgG negative subjects express lower levels of the β-NGF receptor TrkA

Noting that circulating levels of the serum cytokine β-NGF positively associated with the production of anti-NP IgG, we further investigated the implications of this pathway in this setting. β-NGF has been shown to be important in other respiratory viruses, such as RSV (26,27). It is produced by T cells and involved in an induction loop with its receptor Tropomyosin receptor kinase A (TrkA). In line with this, we studied TrkA expression on CD4+ and CD8+ T cells by flow cytometry in relation to β-NGF levels with the different cohorts.

Although the expression of TrkA on both CD4+ and CD8+ T cells appeared low, we were able to demonstrate that by MFI of TrkA on CD4+ T cells positively correlated with serum levels of β-NGF in the whole cohort (**Figure 6A**), yet this parallel was not seen for CD8+ T cells (Figure 6B). The percentage of CD4+ and CD8+ T cells expressing the β-NGF related receptor TrkA was similar between the groups (**Figure 6C and 6D**); however, CD4+ T cells from anti-NP IgG positive subjects express higher levels of TrkA on their surface, when analysed by median fluorescence intensity (MFI) (**Figure 6E**). The same discrepancy, however, was not identified for CD8+ T cells (**Figure 6F**), suggesting a preferential involvement of CD4+ T cells in this β-NGF/TrkA signalling pathway, implying the potential for CD4+ T cell help in this setting.

**Figure 6.**
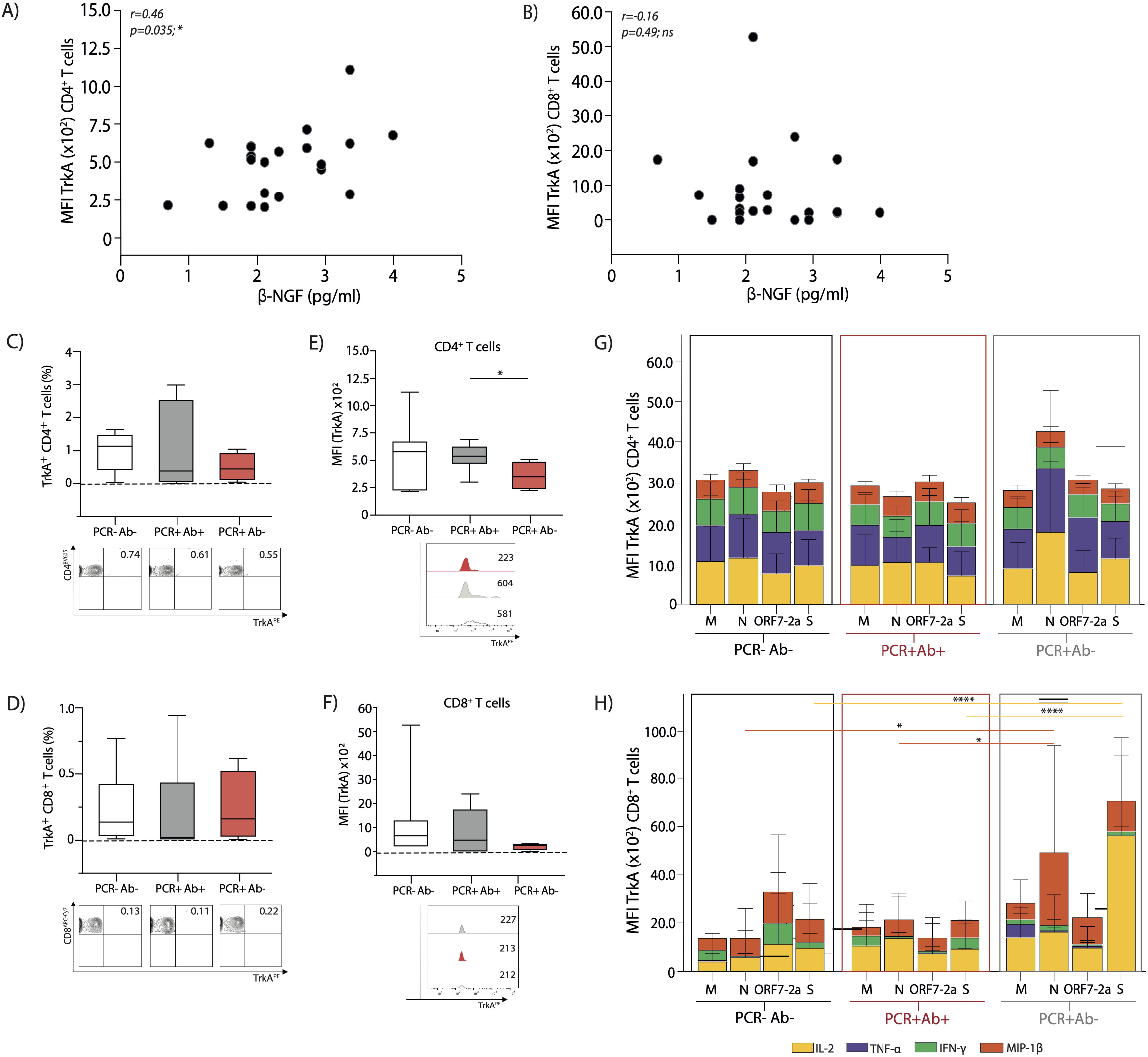
Global and antigen specific T cell expression of the β-NGF receptor TrkA, from the different cohorts. Correlative expression of β-NGF against MFI of TrkA^+^ **(A)** CD4^+^ and **(B)** CD8^+^ T cells from all subjects studied. Summary data of percentage TrkA^+^ **(C)** CD4^+^ and **(D)** CD8^+^ T cells (top), with representative FACS plots (bottom) from each cohort. Summary data of MFI expression of TrkA on **(E)** CD4^+^ and **(F)** CD8^+^ T cells (top); with representative MFI histograms from each cohort (bottom). MFI of TrkA^+^ **(G)** CD4^+^ and **(H)** CD8^+^ T cells producing the respective cytokines after 16-hour stimulation with the four peptide pools derived from SARS-CoV-2. A Spearman non-parametric correlation test was undertaken to test significance in (A) and (B), a one-ANOVA (C-F) and a two-way ANOVA (G, H) with a Tukey’s post-hoc test for multiple comparisons was used to demonstrate significance. Coloured lines for significance indicate changes relative to the corresponding cytokine. *p<0.05; **p<0.01; ***p<0.001; ****p<0.0001; ns - not significant. (PCR-Ab- n=9; PCR+Ab+ n=8; PCR+Ab- n=4).

We then compared TrkA expression (by MFI) on antigen specific CD4+ TrkA+ and CD8+ TrkA+ T cells, to establish if there were discrepancies in relation to SARS-CoV-2 peptide specificities and this signalling pathway. We did not detect any differences in cytokine producing cells between groups with regards to antigen specific CD4+ TrkA+ cells, disparate to the findings on global CD4+ T cells (**Figure 6G**). Overall, cytokine production from CD8+ T cells (by MFI) remained very low, and although differences were detected between both peptide specificities and groups, we would interpret these findings with caution (**Figure 6H**).

## Discussion

Despite the success of the vaccine against SARS-CoV-2 in preventing severe COVID-19, there remains a substantial burden of COVID-19 on healthcare services globally. A deeper understanding of the immune response to SARS-CoV-2 infection and of its inter-individual variability is of a great importance for the implementation of further vaccination strategies during the second year of the pandemic and in the forthcoming years. There has been great progress made in the understanding of the host-virus interaction and the pathogenesis of SARS-CoV-2, mostly limited to severe cases. However, as it was already suggested by early reports (28), SARS-CoV-2 infection leads to mild disease in the majority of cases. An extensive characterisation of the functionality and durability of the immune response in subjects with mild COVID-19 will be instrumental for an in depth risk evaluation and more efficient utilisation of treatment and prophylaxis strategies.

In this work, we analysed the SARS-CoV-2 specific T cell response and the cytokine profile of 17 mild COVID-19 convalescent subjects (positive PCR test) and 10 unexposed subjects (negative PCR test, no symptoms) from a previously published cohort (2), to determine host factors influencing primarily the production of anti-NP antibodies, but also neutralising antibodies. Our initial observation was that seven out of eighty (8.75%) PCR confirmed subjects in our cohort had undetectable anti-NP IgG, in line with other published cohorts where 1-10% of subject did not seroconvert (4,10–12).

We analysed the T cell response by IFN-γ ELISPOT assay. Interesting we noted differences between anti-NP IgG positive and anti-NP IgG negative convalescent patients, where none of the anti-NP negative subjects were able to produce IFN-γ after stimulation with peptides derived from the S protein. Of note, two out of the four analysed anti-NP IgG negative subjects had a detectable IFN-γ response after overnight incubation with peptides derived from NP. This is not surprising, since a SARS-CoV-2 specific T cell response has been detected in mild COVID-19 convalescent subjects even in the absence of seroconversion (29), and the T cell response rather than humoral response is considered to have a determining role in viral clearance (30) as recently shown in rapid abortive SARS-CoV-2 infection (31).

While anti-NP IgG are representative of the humoral response, since they are directed against a very abundant viral protein found inside viral particles or infected cells, they are not indicative, per se, of effective immunity, where the considered hallmark is neutralising antibodies. Thus, we determined the titres of nAbs in our cohort, showing that the IC50 positively correlated with anti-NP levels (Architect Index) in the overall cohort. However, five out of seven anti-NP negative subjects showed evidence of neutralising activity, discordant with the Architect Index. A similar discrepancy between antibody presence and T cell response was observed, where anti-NP IgG presence/titre and or nAbs did not correlate with T cell responses. Similar incongruity has been shown in a larger cohort of healthcare workers, where a multi-specific T cell response was not always associated to the presence of nAbs (14).

To further characterise the T cell response in our cohort we analysed the production of four effector cytokines. While the frequencies of antigen-specific T cells were not different between groups, we observed that T cells from anti-NP positive and negative subjects reacted differently to the peptide pools in terms of number of cytokines produced after overnight stimulation. Particularly, CD4+T cells from anti-NP positive subjects reacted preferentially to peptides derived from S (12.5% of them produced four cytokines after stimulation), followed by the pools derived from M, NP and ORF-7a-2. On the contrary, CD4+ T cells from anti-NP negative subjects reacted weakly to the S-derived pool in comparison to the other antigens and reacted preferentially to M-derived peptides followed by the pools derived from NP and ORF-7a-2. CD8+ T cells from anti-NP IgG positive subjects also strongly reacted to the S-derived pool; CD8+ T cells from anti-NP negative subjects responded similarly to the S pool, while reacted weakly to NP-derived peptides. These observations suggest that a differential hierarchy of cytokine response exists among convalescent subjects, in relation to their ability to produce anti-NP antibodies.

The presence of specific antibodies in our cohort does not directly correlate with the detection of the corresponding antigen specific CD4+ T cells. While none of the four anti-NP subjects for whom PBMC were available produced IFN-γ against S in the ELISPOT assay, three produced at least one cytokine against the same pool when analysed by flow cytometry, and all had detectable nAbs. Similar findings were reported by Marklund et al., (4) where all subjects with mild COVID-19, with undetectable anti-NP IgG demonstrated neutralising activity. However, in our case, a direct correlation between these results cannot be deduced since the S-derived peptides used in our experiments do not cover the RBD. In addition, we were unable to study the T cell response of the two anti-NP negative subjects lacking nAbs, since their PBMC were not available.

To further ascertain if soluble circulating factors may govern anti-NP production, we analysed a panel of cytokines related to infection and inflammation from the serum of subjects between groups. Surprisingly, we found that anti-NP positive and anti-NP negative subjects differed in the levels of only two serum cytokines, β-NGF and IL-1α. A linear correlation was also found between Architect Index values and serum levels of GM-CSF, β-NGF, IL-1α, PBEF and IL-12 p70. However, since the relationship between Architect Index and the subject’s IgG concentration is monotonic but not necessarily linear across its range, we decided to consider the presence of anti-NP IgG as a binary variable (either positive or negative), and perform a binary logistic regression, from which only β-NGF levels positively correlated with anti-NP IgG positivity. On the contrary, no correlation was found between β-NGF serum levels and nAbs titres, suggesting that the production of anti-NP and nAbs may be subject to different dynamics and kinetics.

β-NGF is the active form of the first discovered member of a family of neurotrophines (32,33). It is expressed and released by a variety of cell types including T and B lymphocytes (22,23,34); its low basal expression levels increase during inflammation, and it can be induced by pro-inflammatory cytokines such as IL-1β, TNF-α and IL-6 (35,36). The biological effects of β-NGF are mediated by two receptors: p75NTR (low-affinity, can bind other neurotrophines) and Tropomyosin receptor kinase A (TrkA, high affinity and β-NGF-specific) (25,35). Activation of the TrkA receptor leads to cell survival, proliferation, differentiation, and activation. Engagement of the low affinity p75NTR receptor in the absence of TrkA activates the pro-apoptotic pathway. Basal expression of TrkA is up-regulated on B and T cells after antigenic or inflammatory stimulation; moreover, in some cell types, expression or circulating levels of β-NGF strongly correlate with TrkA expression, suggestive of a positive feedback loop (37,38). Since the high-affinity TrkA receptor is expressed on human CD4+ T cells (24), we determined its expression by flow cytometry on T cells isolated from subjects in our cohort. We observed that the frequency of CD4+ TrkA+ T cells was similar in all three groups, but the median fluorescence intensity (MFI), indicative of the quantity of TrkA molecules expressed on the surface of the cells, was lower in the anti-NP negative than in the anti-NP positive subjects. Such difference, however, was not confirmed in antigen specific CD4+ T cells, where high variability within groups was observed. β-NGF serum levels positively correlated with TrkA expression on global CD4+ T cells, as expected based on the suggested positive feedback loop between the cytokine and its receptor.

Due to the cross-sectional nature of our study, we are not able to determine whether the β-NGF levels observed are either a consequence of the recent SARS-CoV-2 infection or reflect the basal levels of our subjects. However, since β-NGF basal levels are normally low, it is possible that the lower plasma levels observed in the anti-NP negative subjects reflect an attenuated inflammatory response experienced by these subjects during the infection; this would be in line with the more severe presentation of the disease in patients with an anti-NP dominated humoral response (5,20,21).

Interestingly, β-NGF has been studied in the contexts of respiratory syncytial virus (RSV), and human rhinovirus (HRV) infection, where it inhibits apoptosis of the bronchial epithelial cells supporting viral replication, and promotes virus internalisation, respectively (39,40). To our knowledge, an involvement of β-NGF in SARS-CoV-2 infection has not been investigated, but our results indicate a potential association between the β-NGF/TrkA signalling pathway and the production of anti-NP antibodies, which in turn reflects a different degree of inflammation caused by SARS-CoV-2 infection.

We acknowledge the limitations of our study, in part linked to the difficulties in diagnosis during the early phase of the pandemic, when PCR testing was not widely available. For this reason, we were not able to determine the exact timeframe between symptom onset, PCR test and sample collection for most of our subjects. Moreover, our sample size was limited, especially the group of PCR positive anti-NP negative subjects; this could be considered an intrinsic limitation of this research field, since only 1-10% of PCR confirmed cases are estimated to have undetectable antibodies. In addition, we acknowledge the fact that only four out of seven anti-NP negative, PCR positive subjects in our cohort had given their consent for PBMC isolation from venous blood. The ex-vivo assays were not performed using overlapping peptides covering the entire sequence of the SARS-CoV-2 proteins, but only selected peptides were used; however, since they were found to be immunogenic (17,27) we are confident that the results we obtained are representative, and are in line with other studies reviewed by Bertoletti et al.(30).

We describe for the first time the β-NGF/TrkA signalling pathway as a host factor reflecting different levels of inflammation within mild COVID-19 cases, with effects on the virus-specific humoral and T cell response. The mechanistic regulation of this pathway in COVID-19 disease deserves further investigation, and larger studies are required to determine whether the effects of such differences can influence the durability of the T cell response and vaccine-induced immunity.

## Data Availability

All data produced in the present study are available upon reasonable request to the authors

## Author Contributions

CU, USG and PTFK designed the research study; CU, JMG, CP and USG conducted the experiments; CU, JMG, CP and USG acquired data; CU, WL and SRJ, AMK, USG analysed data; AMK, USG and PTFK provided reagents; CU drafted the manuscript; all authors critically revised the manuscript and approved the final version.

## Acknowledgments

The authors would like to thank Dr Louisa James (Blizard Institute, Barts and The London SMD, QMUL), Dr Sefina Arif and Norkhairin Yusuf (King’s College London) for technical assistance. We would also like to thank all clinical and administrative staff at The Graham Hayton Unit, Royal London Hospital for aiding with recruitment and clinical sample collection.

