## Supplementary table and figures for "The β-NGF/TrkA signalling pathway is associated with the production of anti- nucleoprotein IgG in convalescent COVID-19"

**SUPPLEMENTARY DATA**

| **PCR** | **Anti-NP** | **N** | **Ethnic composition (Asian; Black; White)** | **Age (median; IQR)** | **% of females** |
| --- | --- | --- | --- | --- | --- |
| - | - | 10 | 4; 0; 6 | 37; 42.25-30 | 50 |
| + | + | 10 | 3; 0; 7 | 38; 51.5-29.25 | 50 |
| + | - | 7 | 4; 0; 3 | 34; 36.5-27.5 | 42.9 |
| N/A | + | 13 | 3; 1; 9 | 40; 48-34 | 61.54 |
| p-value | | | | ns | ns |

**Supplementary Table 1. Demographic characteristics of the 40 selected participants; p-values lower than 0.05 were considered significant. ns= not significant**

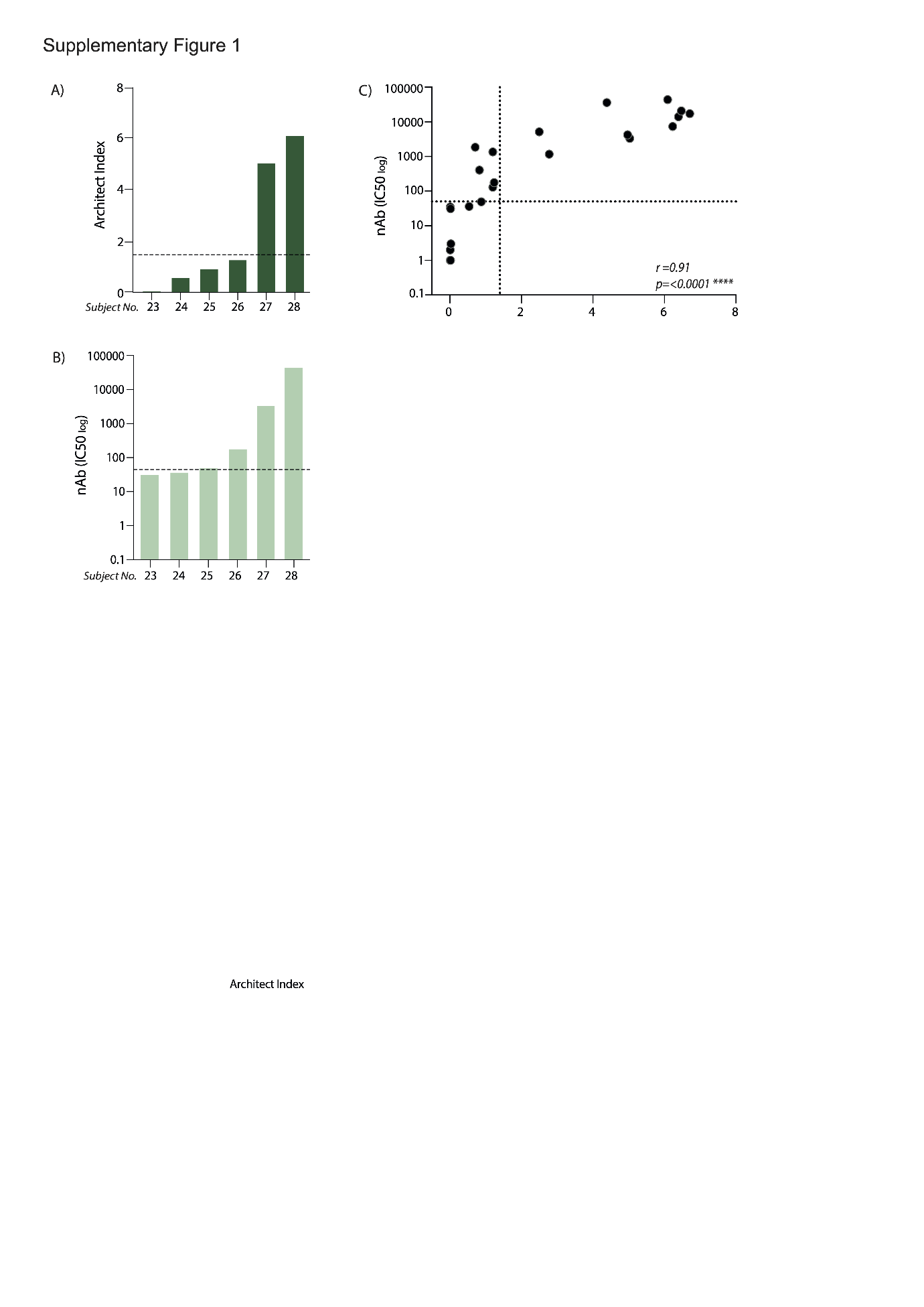

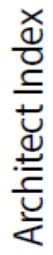

**Supplementary Figure 1; Comparison between anti-NP levels and nAb titres. (A)** anti-NP IgG levels expressed as Architect Index (manufacturer arbitrary units) for subject analysed in the study for which T cell ELISpot analysis was not undertaken (ordered lowest to highest); the dotted line represents the 1.4 cut-off, below which samples are considered as negatives. **(B)** Neutralising antibody (nAb) titres (IC50) corresponding to the same subjects in (A); the dotted line represents the cut-off below which samples are considered as negatives (IC50=50). **(C)** Correlation between anti-NP IgG levels and nAb titres in all subjects (n=28). The dotted lines represent the cut-offs below which samples are considered negatives for each assay. A Spearman non-parametric correlation test was used to determine significance, *p<0.05; **p<0.01; ***p<0.001, ****p<0.0001, ns = not significant.

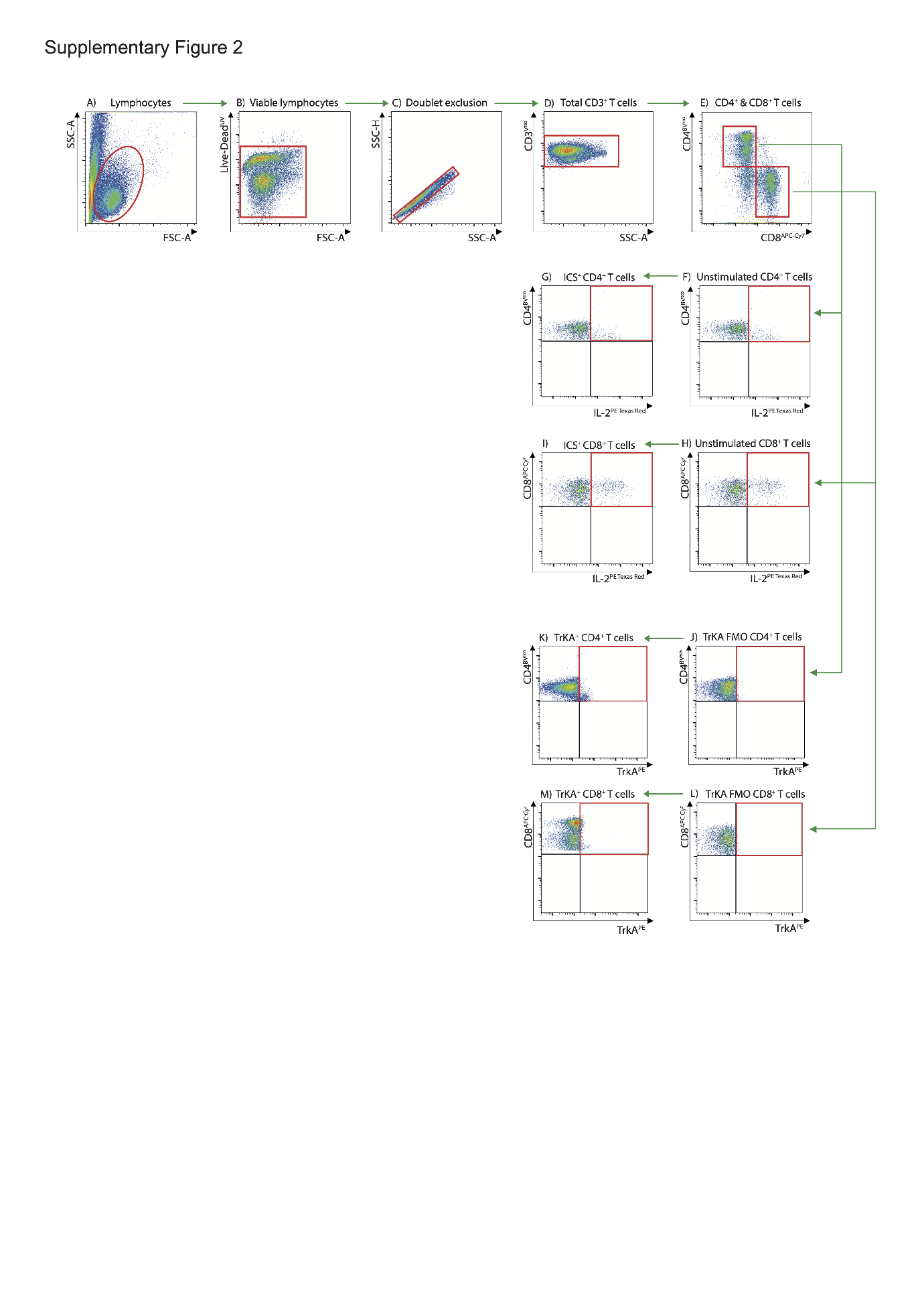
**Supplementary Figure 2**; **Flow cytometry gating strategy.** Initial gating of **(A)** Total lymphocytes with forward scatter (FSC)/side scatter (SSC). To include only **(B)** viable cells for analysis, dead cells were excluded using a viability dye. **(C)** Single cells were then identified defined on an SSC-A/SSC-H plot. **(D)** Cells were then gated on the CD3^+^ population, and further gated on the **(E)** CD4^+^ and CD8^+^ subpopulations. Antigen-specific CD4^+^ and CD8^+^ cells were gated according to the cytokines produced, **(F)** unstimulated CD4^+^ T cells and **(G)** intracellular cytokine staining (ICS) staining of CD4^+^ T cells following peptide stimulation (IL-2 shown), in addition **(H)** unstimulated CD8^+^ T cells and **(I)** ICS staining of CD8^+^ T cells following peptide stimulation (IL-2 shown). TrkA analysis, by **(J)** FMO on CD4^+^ T cells and **(K)** with TrkA strained CD4^+^ T cells, in addition **(L)** FMO on CD8^+^ T cells and **(M)** with TrkA strained CD8^+^ T cells.

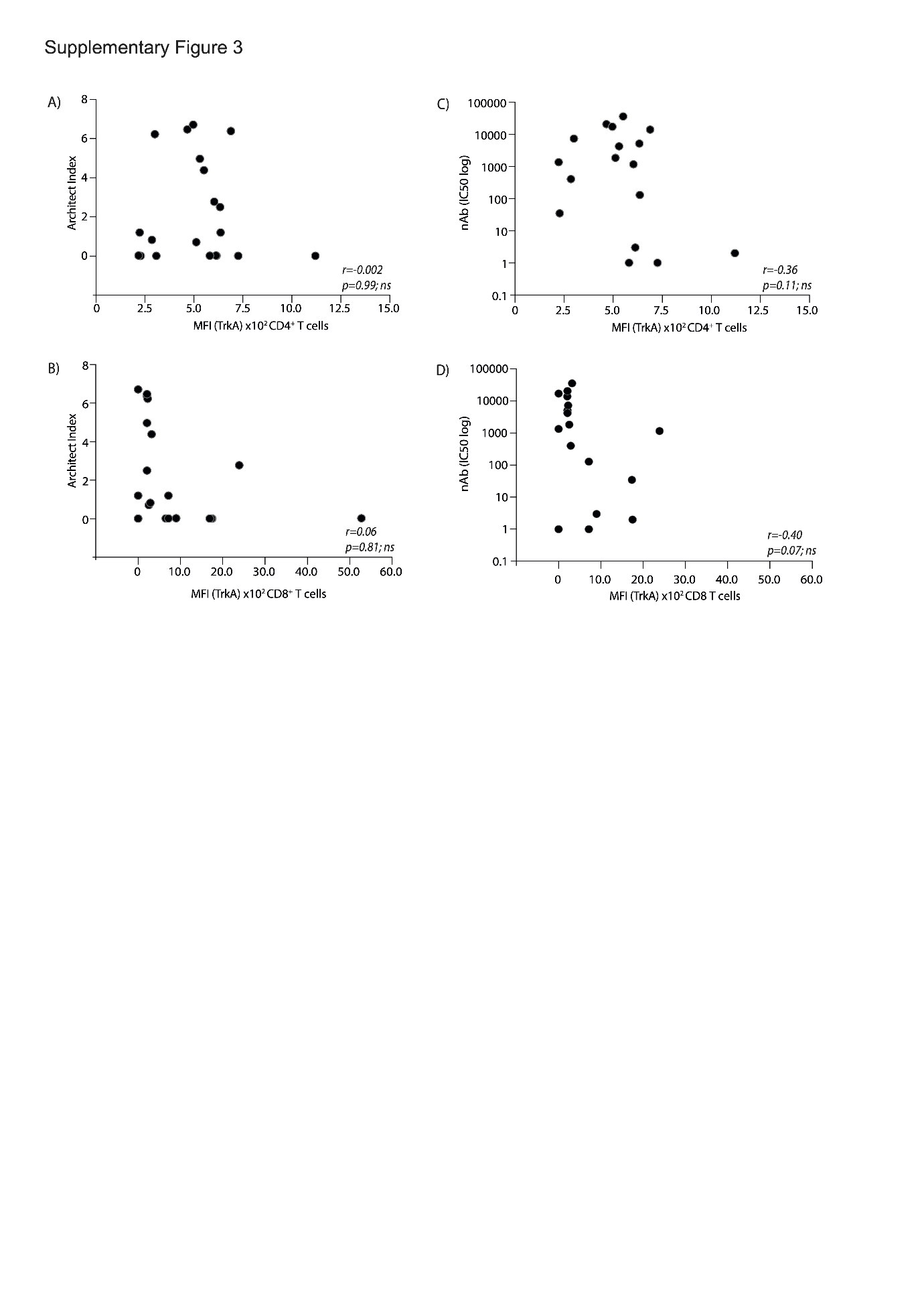

**Supplementary Figure 3; TrKA^+^ T cells in relation to antibody production.** Correlative expression of anti-NP levels (Architect index) in relation to TrKA^+^ expressing **(A)** CD4^+^ and **(B)** CD8^+^ T cells along with nAb levels in relation to TrKA^+^ expressing **(C)** CD4^+^ and **(D)** CD8^+^ T cells. Spearman non-parametric correlation tests were used to determine significance, *p<0.05; **p<0.01; ***p<0.001, ****p<0.0001, ns = not significant.
